# Ongoing outbreak of COVID-19 in Iran: challenges and signs of concern with under-reporting of prevalence and deaths

**DOI:** 10.1101/2020.04.18.20070904

**Authors:** Mahan Ghafari, Bardia Hejazi, Arman Karshenas, Stefan Dascalu, Alireza Kadvidar, Mohammad A. Khosravi, Maryam Abbasalipour, Majid Heydari, Sirous Zeinali, Luca Ferretti, Alice Ledda, Aris Katzourakis

**Author notes:** Corresponding author. (MG); (AK).

## Abstract

Many countries with an early outbreak of SARS-CoV-2 struggled to gauge the size and start date of the epidemic mainly due to limited testing capacities and a large proportion of undetected asymptomatic and mild infections. Iran was among the first countries with a major outbreak outside China. Using all genomic sequences collected from patients with a travel link to Iran, we estimate that the epidemic started on 21/01/2020 (95% HPD: 05/12/2019 – 14/02/2020) with a doubling time of 3 days (95% HPD: 1.68 – 16.27). We also show, using air travel data from confirmed exported cases, that from late February to early March the number of active cases across the country were more than a hundred times higher than the reported cases at the time. A detailed province-level analysis of all-cause mortality shows 20,718 (CI 95%: 18,859 – 22,576) excess deaths during winter and spring 2020 compared to previous years, almost twice the number of reported COVID-19-related deaths at the time. Correcting for under-reporting of prevalence and deaths, we use an SEIR model to reconstruct the outbreak dynamics in Iran. Our model forecasted the second epidemic peak and suggests that by 14/07/2020 a total of 9M (CI 95%: 118K – 44M) have recovered from the disease across the country. These findings have profound implications for assessing the stage of the epidemic in Iran and shed light on the dynamics of SARS-CoV-2 transmissions in Iran and central Asia despite significant levels of under-reporting.

**One Sentence Summary:** We use epidemiological and genetic data to investigate the origins of the SARS-CoV-2 outbreak in Iran and assess the degree of under-reporting in prevalence and deaths.

## Introduction

With significant levels of mortality and morbidity, the ongoing pandemic of Coronavirus Disease 2019 (COVID-19) continues to have a major impact on many countries around the world [1]. First reported cases of COVID-19 in Iran were of two infected individuals in the province of Qom on 19 February. In only a few days, reported cases grew rapidly in almost a dozen provinces (see Movie S1 and Figure 1a) which raised concerns that the virus had already been widespread by the time the first two cases were detected in Qom. By 28 February, WHO reported nearly 100 confirmed exported cases of COVID-19 from Iran to several countries [2] while, at the time, the Ministry of Health and Medical Education (MoHME) reported a total of 388 confirmed cases across the country [3]. Some early reports suggested that the first two confirmed cases in Qom, one of the thirty-one provinces of Iran, could have been infected by a merchant who had reportedly travelled from China [4]. However, with COVID-19, identifying the so-called ‘patient zero’ is problematic due to high rates of asymptomatic and pauci-symptomatic infections [5]. Indeed, some preliminary analysis by the COG-UK consortium shows that there have likely been more than one thousand unique introductions of the virus into the UK, possibly many of which are from individuals who are asymptomatic or show mild symptoms [6]. Therefore, the most likely route of virus spread from China to Iran, and indeed many other countries, was via asymptomatic travelers. Infected individuals typically show no symptoms for about 5 days [7-9] or sometimes no symptoms at all [10-12], while silently spreading the virus [14]. Using aggregate data from the influenza surveillance networks has been proposed as one of the ways to detect early arrival of SARS-CoV-2 into a country [13]. However, a major flu outbreak in Iran which reportedly stretched from November 2019 to early January 2020 may have conflicted with the early diagnosis of COVID-19 patient across the country, thereby hindering control and diagnosis efforts [15,16].

**Fig. 1.**
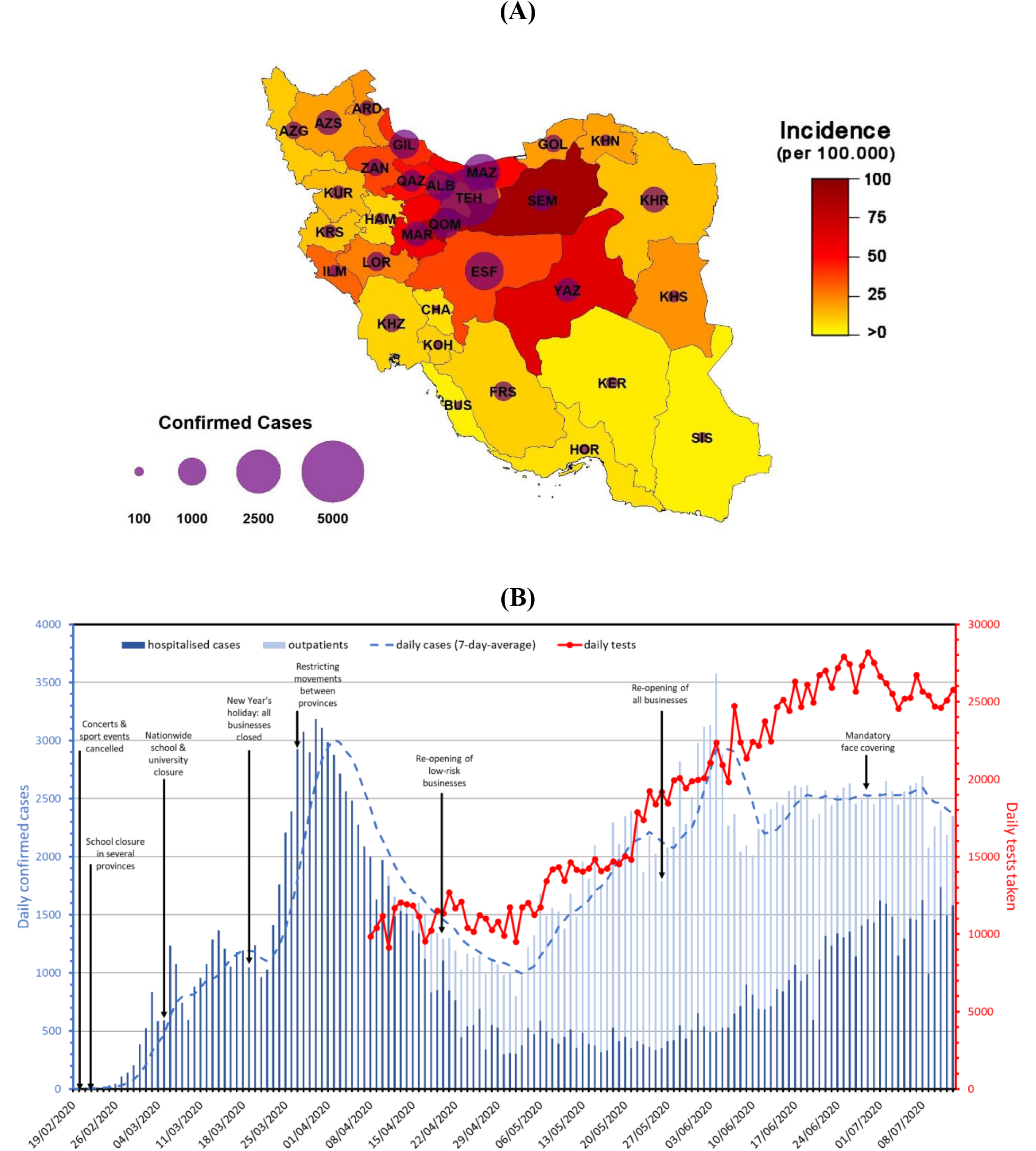
Epidemiological and demographic characteristics of confirmed COVID-19 cases in Iran through time. (**A**) Total incidence of COVID-19 by province on 22 Mar 2020 (last day that MoHME released province-level daily cases). (**B**) Number of confirmed hospitalised patients (dark blue) and outpatients (light blue). Red lines show the number of daily tests taken over time. Vertical arrows indicate some of the major interventions or changes of policies.

During the early days of the epidemic, Pasteur Institute of Iran (PI) was the primary source of testing suspect COVID-19 cases and had enough resources to perform only a few hundred RT-PCR tests per day. By mid-March, WHO and the Chinese Government delivered several shipments of emergency medical supplies along with more than 200,000 additional test kits which helped with the diagnosis of more hospitalised patients [17]. Nevertheless, some of the preliminary analysis showed that Iran was only reporting less than 10% of its symptomatic cases during its first peak in late March [18,19]. From early April, as the country ramped up its testing capacity, MoHME started to report outpatients and carried out limited levels of contact tracing in several provinces (see Fig. 1b). However, despite the growing testing capacity, since the government announced plans for re-opening high-risk activities in late May, the second peak in new cases emerged and the number of outpatients started to shrink over time. This raised concerns that, like the first peak when hospitals in many provinces were at near maximum capacity, the degree of under-reporting in both prevalence and deaths has raised substantially higher again. Another point of concern is that the rate of false-negative detection of SARS-CoV-2 by RT-PCR tests can get significantly higher over time as the delay from the onset of symptoms to testing increases. Therefore, by the time tests are taken from hospitalised cases of COVID-19, there could be typically 3 to 7 days past since the onset of their symptoms and, as a result, a significant portion of their tests (up to 50%) may come out as negative [7,20].

In this study, we provide a detailed analysis of the COVID-19 outbreak in Iran. By gathering genetic and air travel data from passengers with a link to Iran, we estimate the start date of the epidemic and its prevalence across the country. Coupling this with the estimated number of excess deaths and other key clinical and epidemiological information, such as the age-stratified infection fatality ratios and delays from onset of infection to death, enables us to reconstruct the full transmission dynamics of COVID-19 despite significant levels of under-reporting in prevalence and deaths.

First, we investigate the possibility of data manipulation on reported number of cases and deaths by MoHME using Benford’s law. We then use epidemiological and genetic data from air travelers with a travel history to Iran along with the first whole-genome sample obtained from inside the country to determine the start date and early growth rate of the epidemic. Next, we provide a province-level analysis on the pattern of spread during winter and spring, highlighting the degree of under-reporting in both point prevalence and deaths. We also assess the risk of importation of infected individuals from China into the country from January to mid-March 2020. Finally, we evaluate the impact of non-pharmaceutical interventions (NPIs) on the growth rate of the epidemic and use an SEIR model to evaluate its burden on the healthcare system over the course of the outbreak.

### Results

### Investigating data manipulations using Benford’s law

By comparing the reported prevalence and deaths in several countries with sizable outbreaks from mid-February to early-April, we find that many countries, including Iran, were initially on an exponential growth trajectory (see Fig. 2a and 2b). Depending on the testing strategy of each country, i.e. whether they only test hospitalised patients or also allow testing for outpatients, we would expect the reported death toll to follow the same trend as confirmed cases with a few days of delay. However, since 22 March, after passing a total of 1,000 confirmed deaths, Iran’s reported death toll changed course to a linear growth despite the fact that the number of cases were still growing exponentially up to 2 April. This appeared in stark contrast with many other countries with large outbreaks and raised concerns over the credibility of the reports from Iran and possible manipulation of data by MoHME. We investigate the latter using Benford’s Law (see Methods section). Fig. 2c shows the distribution of leading digits in reported cases and deaths during the exponential phase of the outbreak (see Fig. 2a and 2b) for Iran, USA, and UK which we compare to the Benford distribution. The result shows that the distribution of leading digits in all the three countries are similar to Benford’s distribution and that there is no evidence to suggest a manipulation of data has occurred in any of the examined data sets. It is likely that the apparent discrepancy in MoHME’s reported numbers is due to delayed turnaround times at the peak of the outbreak when hospitals in several provinces were at near maximum capacity. Prioritising testing for active cases over post-mortem testing of suspect cases is also likely to have contributed to the observed difference in trends.

**Fig. 2.**
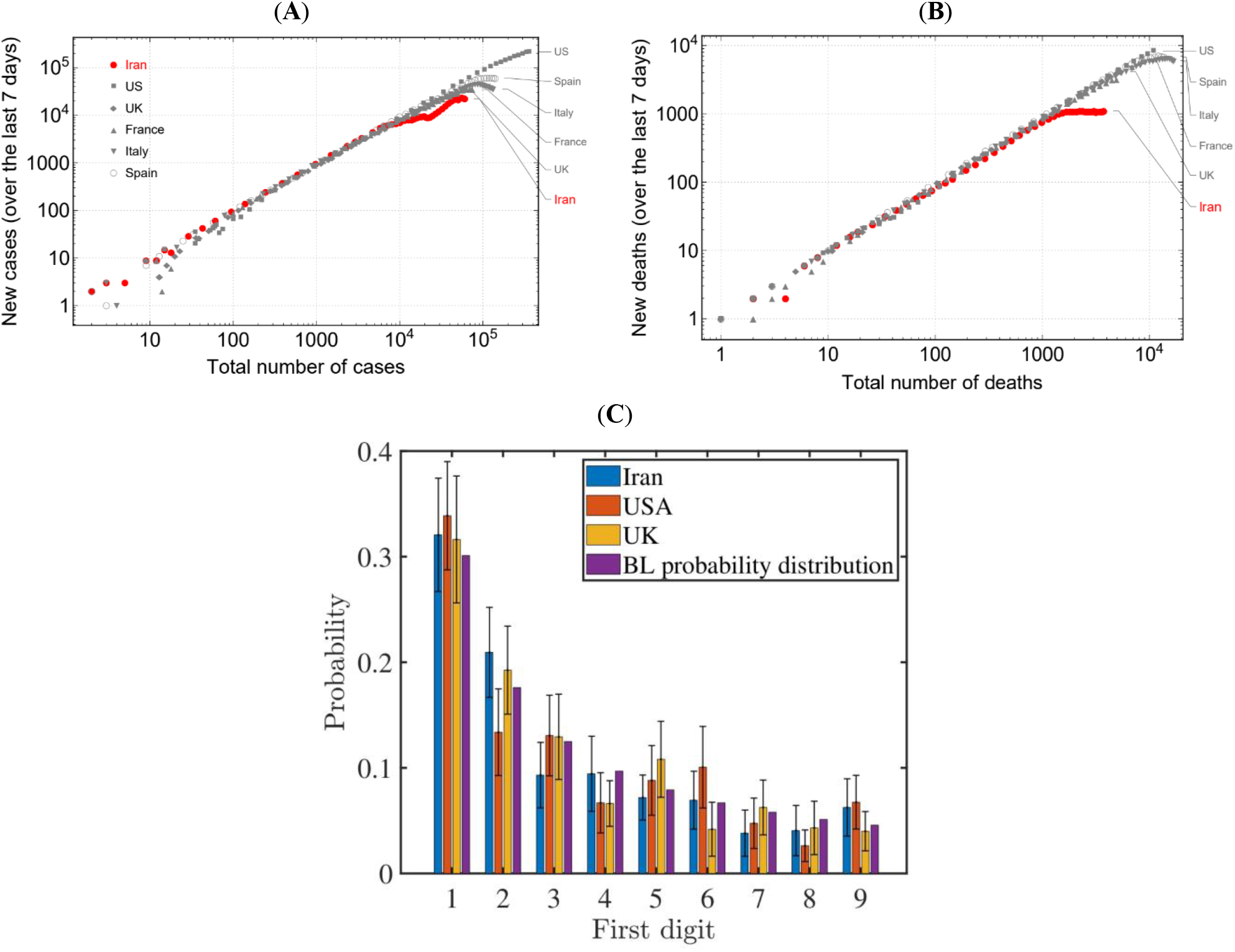
Investigating data manipulation on reported number of cases and deaths. (**A**) and (**B**) show the number of new cases and deaths over the last seven days with respect to the total number of cases and deaths, respectively. (**C**) Shows the probability distribution of leading digits in the total number of confirmed cases and deaths from the start of the epidemic in each country during the exponentially growing phase and compare this to the distribution given by Benford’s law (BL). We sample 40 random numbers from this data set for each country and count the number of occurrences for the leading digits. To ensure convergence, we repeat this process 50 times. Error bars represent one standard deviation unit away from the mean.

### Origins of the epidemic in Iran: a phylogenetic analysis

Since the virus spread undetected during the initial phase of the Iranian epidemic, no epidemiological information is available for this phase. Molecular epidemiology provides a powerful tool to reconstruct the early epidemic trajectory a posteriori, especially when direct epidemiological data are absent, incomplete or biased. The 16 March report from NextStrain showed that sequences from cases with a travel history to Iran correspond to a distinct clade of SARS-CoV-2 [21] and that they were phylogenetically related to the dominant variant circulating in Wuhan at the time (prototype strain MN908947/SARS-CoV-2/Wuhan-Hu-1) [22] – see Data S1 for the full list of genome metadata used for this analysis. We first construct a maximum likelihood phylogenetic tree of all sequences that were linked to Iran (see Fig. 3a). We then use these genomic sequences to determine the time to the most recent common ancestor (TMRCA) of the sequences and characterise the initial epidemic growth rate in Iran (see Methods section). Consistent with other studies [23,24], our inferred substitution rate is 1.66 × 10^−3^ (95% Highest Posterior Density (HPD): 2.18 × 10^−4^ − 3.31 × 10^−3^) per site per year and the exponential growth rate of 82 in units of years, corresponding to a doubling time of around 3 days (95% HPD:1.68 – 16.27), which is close to the growth rate in the initial phase of European COVID-19 epidemics [56]. The age of the root is placed on 21/01/2020 (95% HPD: 05/12/2019 – 14/02/2020), a month earlier than the first reported case.

**Fig. 3.**
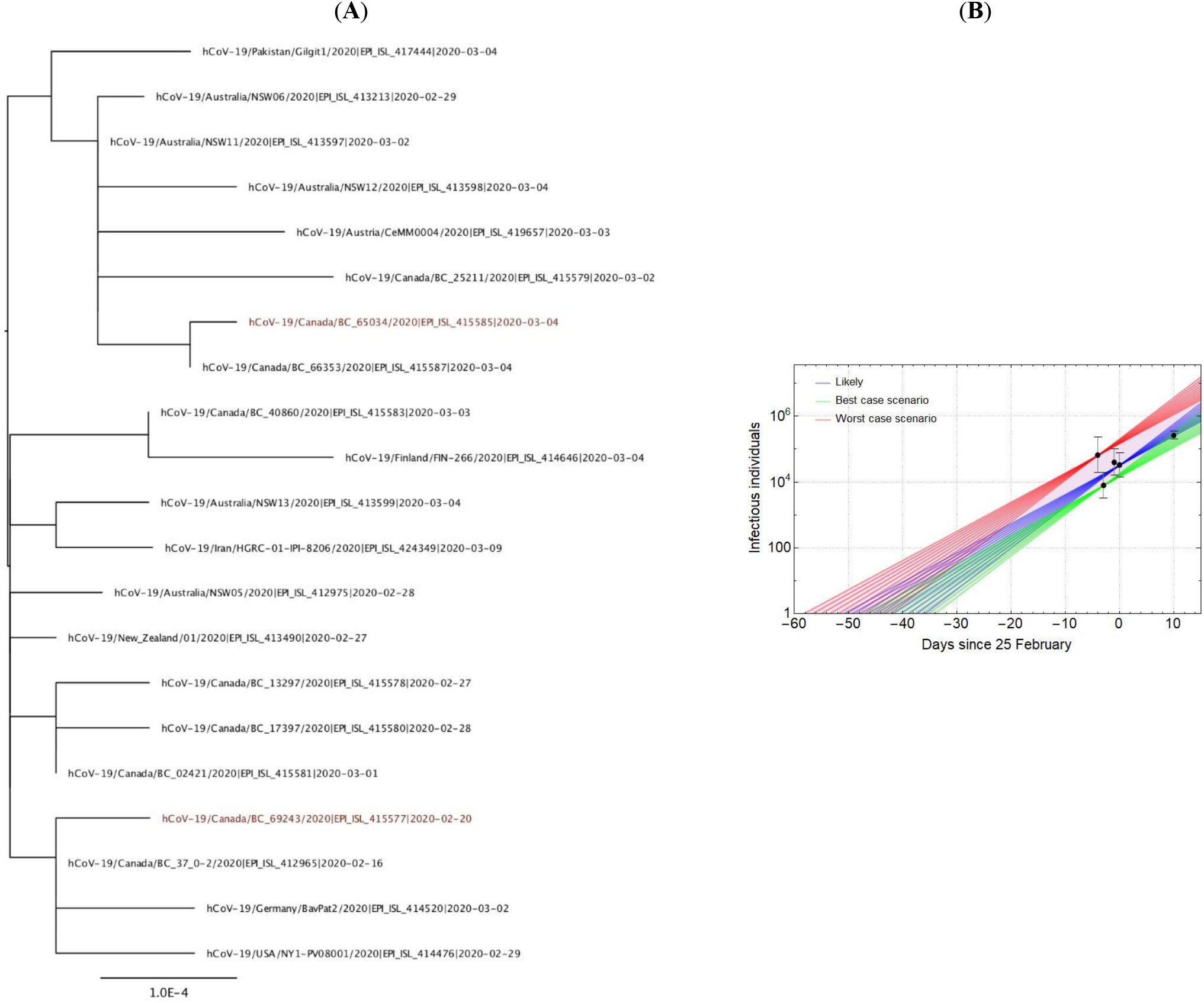
Phylogenetic and epidemiological analysis on cases with a travel link to Iran. (**A**) Maximum likelihood phylogenetic tree of samples linked to Iran (including the first sample from inside Iran, HGRC-01-IPI-8206/2020). The labels include the sampling times used in the BEAST analysis. Red labels indicate the two epidemiologically linked samples that were excluded from the subsequent analysis. (**B**) Estimating the start date of the epidemic and its initial growth trajectory (with a range of doubling times from 2.5 to 3.5 days) based on the likelihood analysis on exported cases to UAE (best case scenario with the lowest estimated number of active cases), Lebanon (worst case scenario with the highest estimated number of active cases), and Kuwait, Oman, and China (see Table 1). The likely scenario is based on the expected number of active cases in late February.

### Estimating the under-reporting of incidence using air-travel data

High levels of under-reporting have been noticed in several countries during the early phase of the COVID-19 pandemic. Such under-reporting creates biases that hinders direct estimates of the actual incidence of the disease and are likely to be present in MoHME’s report as well. A more reliable estimate of the size of the Iranian epidemic can be obtained from the number of exported cases detected abroad [57575757]. In late February, a total of 8 internationally exported cases of COVID-19 with direct flights from Iran were detected in Lebanon, UAE, Oman, and Kuwait. Similarly, in early March, an additional 28 cases were identified in China (see Table 1). By finding the approximate number of passengers per week travelling from Iran to these countries, we use a binomial likelihood estimation method (see Methods section) to estimate the nationwide incidence (see Fig. 3b). Our result suggests the number of infectious individuals by 25 February is 18,500 (95% CI: 11,400 – 30,700) and 259,500 (95% CI: 199,200 – 356,200) 6 March which is aligned with estimated outbreak size from other studies [25,26] and is more than 100 times higher than confirmed cases that MoHME reported at the time – see the sensitivity analysis on the parameters of our model in Table S2. These estimates also suggest a doubling time of 2.6 days (approximate 95% CI: 2.2-3.3 days), which is remarkably consistent with the range observed in many other countries with major outbreaks (2.5 – 3.5 days). We then use this range of doubling times to extrapolate the number of cases back in time and find 13/01/2020 (05/01/2020 – 25/01/2020) to be the approximate start date of the epidemic, which is also well aligned with the estimated root age of our phylogenetic analysis.

**Table 1.** Estimated number of infectious individuals based on air travel data. This includes a list of all countries with confirmed cases of COVID-19 travelling from Iran via direct flights (see Data S3 for more information).

| Date | Country | Passengers/week | Reported cases | Estimated active cases (95% CI) |
| --- | --- | --- | --- | --- |
| 21/02/2020 | Lebanon<sup>1</sup> | 800 | 1 | 64,000 (19,700 – 236,900) |
| 22/02/2020 | UAE<sup>2</sup> | 13,430 | 2 | 7,700 (3,200 – 19,900) |
| 24/02/2020 | Oman<sup>3</sup> | 2,660 | 2 | 39,000 (16,100 – 101,000) |
| 25/02/2020 | Kuwait<sup>4</sup> | 4,025 | 3 | 32,200 (14,400 – 76,000) |
| 06/03/2020 | China<sup>5</sup> | 4,500 | 28 | 259,500 (199,200 – 356,200) |

<sup>1</sup>Source, <sup>2</sup>Source, <sup>3</sup>Source, <sup>4</sup>Source, <sup>5</sup>Source

### Estimating the under-reporting of cumulative deaths using excess mortality data

Given the high levels of under-reporting, excess mortality data represent a more reliable source to estimate COVID-19 related deaths. By analysing the seasonal reports from the National Organization for Civil Registration (NOCR) of Iran on province-level all-cause deaths, we find that four provinces in winter and twenty-six provinces in spring 2020 had significantly higher recorded deaths compared to previous years (see Fig.4a and 4b). During winter, the four provinces in central and northern Iran (Qom, Gilan, Golestan, and Mazandaran) showed a 27% rise in mortality with 3,476 (95% CI: 3,172 – 3,779) deaths in excess of previous years. In spring, our estimates show a 22% increase with a total of 17,242 (95% CI: 15,687 – 18,797) excess deaths in twenty-six provinces compared to previous years. Qom and Gilan, two of the hardest-hit provinces in winter, show a lower percentage of excess deaths in spring possibly indicating that the largest part of their outbreak occurred back in winter while in many other provinces such as Mazandaran, Khuzestan, and Qazvin the excess deaths continued to increase during spring. This is also corroborated by numerous reports from various provinces across the country and is aligned with the fact that the first nationwide epidemic peak occurred in late-March.

**Fig. 4.**
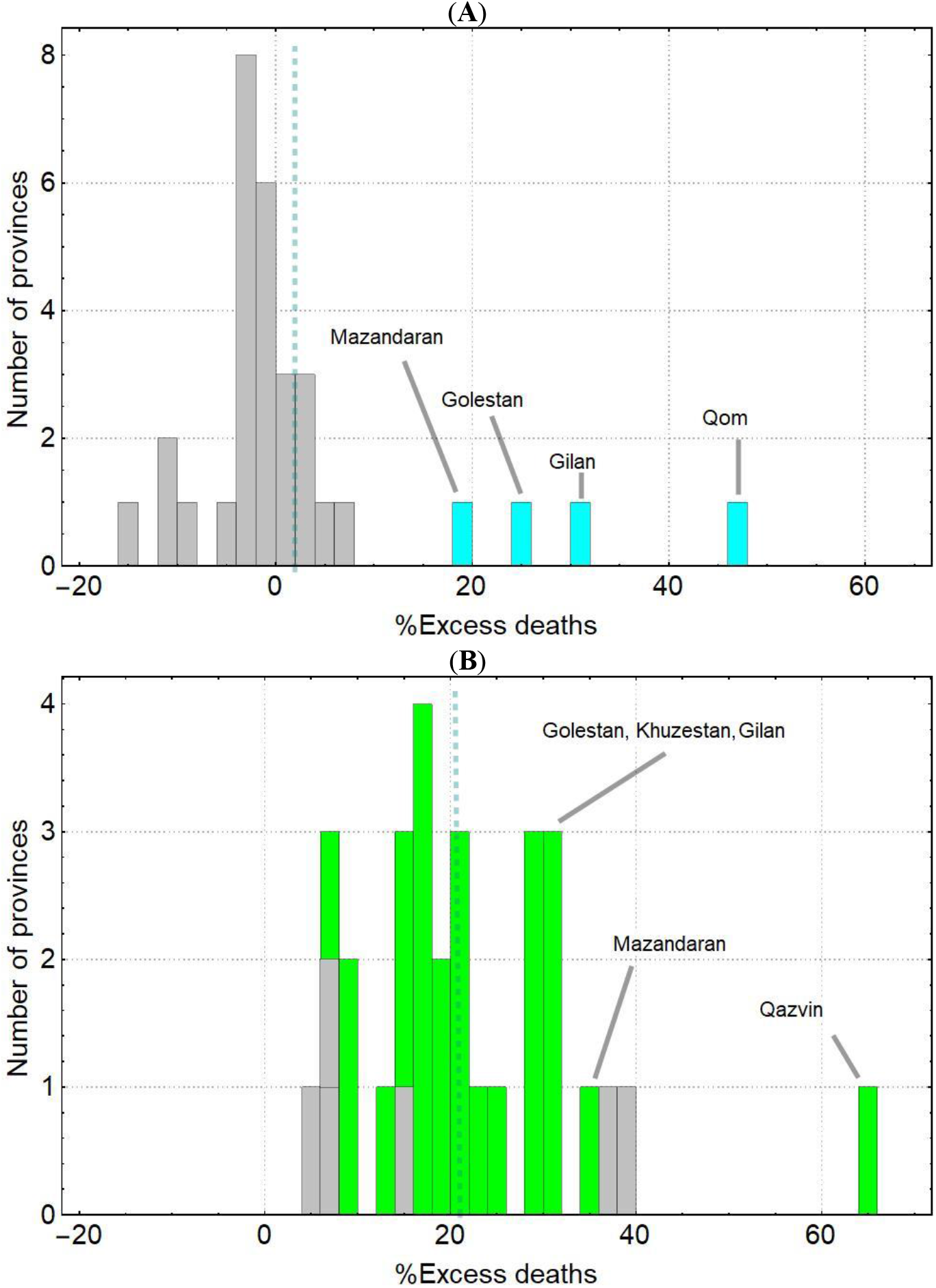
Percentage of excess deaths (with respect to the five-year average) during winter and spring 2020 in 31 provinces of Iran. (**A**) and (**B**) show the percentage of excess deaths during winter (from 22 Dec 2019 to 19 Mar 2020) and spring (from 20 Mar to 20 June 2020). Four provinces (highlighted in cyan) during winter and twenty-six provinces (highlighted in green) during spring show significant levels of excess mortality. Gray bars represent provinces with no significant deviations from their five-year average (based on 95% confidence interval). The vertical dashed lines show the mean percentage of excess mortality in each season. During winter, the mean excess mortality is only 2% while in spring this grows to 21% across all provinces.

### Early-stage importation risk from China

The Iranian epidemic was most likely seeded by one or more imported cases flying from China. Given the debate on travel restrictions, it is worth assessing what was the actual early-stage risk of imported cases in Iran. We match the air travel data from infected passengers travelling via flights from Beijing, Shanghai, and Guangdong with the prevalence data from China (see Methods section) and find that the expected number of imported cases to Iran from 22 January to 15 March remained very low, with just one imported case expected across the whole period. The fact that the epidemic took over despite the low number of importations from China and the strong overdispersion in transmissions suggests that infected individuals likely belong to a high-risk sub-group of the population, such as businessmen, politicians, or other individuals who frequently travel to the country and tend to meet a large number of contacts.

### Evaluating effectiveness of intervention measures and the dynamics of the outbreak

We first evaluate the effectiveness of key NPIs (see Methods section) implemented on set dates in Iran and their impact on reducing the effective reproduction number, Rt (Table 2 and Figure 5a). Our analyses indicate that while intervention measures, particularly the nationwide lockdown during the New Year’s holiday month, were effective in curbing the spread (Rt < 1), the reopening of high-risk businesses in May led to the emergence of a new peak of infection across the country (Rt > 1). They also suggest that mandatory face covering in public spaces can be effective enough to stop the exponential growth pattern (Rt <1) and reduce the transmission rates by late-July/early-August conditioned on having no other major change in NPIs or social behaviour (e.g. travelling for holidays or mass gatherings) during this time period. The model successfully recreates the nationwide trend of the outbreak with one peak in deaths during late March and a second emerging peak that started back in late May due to relaxed NPI measures (see Figure 5b and 5c). Our model correctly forecasted the insurgence of the second epidemic peak [18]. Also, our estimates for point-prevalence and cumulative deaths are aligned with those we predicted based on air travel and excess mortality. Our results suggest, cumulatively, that 9.04 (95% CI: 0.147 – 43.65) million people have recovered and 26.14 (95% CI: 0.53 – 155.22) thousand died as of 14 July 2020. Both the first peak in March and the second peak in July has very likely overwhelmed the healthcare system with approximately 8.70 (95% CI: 0.55 – 85.85) thousand severely ill patients during the peak weeks and 0.474 (95% CI: 0.031 – 4.8) million infectious at the peak of the epidemic (see Data S5). We note the large variation in the results is due to variations in the effective reproduction number, Rt, and its significant impact on the growth dynamics of the epidemic. The upper bound in the simulation results corresponds to an initial reproduction number of Rt = 5 in which case most infections and deaths take place during the first peak in late March and Rt never goes below 1 throughout the simulation (population reaches herd-immunity threshold by mid-June) while the lower bound assumes an initial reproduction number Rt = 2.5 with a very mild first peak in March after which Rt remains below 1 and no second peak will emerge.

**Table 2.** Timeline of intervention policy announcements and their impact on effective reproduction number.

| Intervention measures | Description | Date effective† | Relative %reduction in Rt (range) ‡ | Reference |
| --- | --- | --- | --- | --- |
| School & university closure | Implemented in multiple provinces: Tehran, Mazandaran, Qom, Qazvin, Golestan, Gilan, and several others. | 22/02/2020 | 20%<br>(10-30) | [47] |
| Nationwide lockdown* | Closure of all schools, universities, sport centres, & holy sites. | 05/03/2020 | 80%<br>(65-90) | [47] |
| Re-opening of low-risk businesses | Partial re-opening of certain businesses such as banks and governmental agencies with limited staff. | 18/04/2020 | 70%<br>(60-85) | [47,48] |
| Re-opening of all businesses | Re-opening of all businesses including shops and restaurants while observing social distancing rules | 26/05/2020 | 60%<br>(50-75) | [48,49] |
| Mandatory face-covering | Mandatory in all public spaces including public transports, restaurants, & shops | 05/07/2020 | 75%<br>(60-90) | [53] |

\*This intervention is followed by the closure of all business across the country due to the Iranian New Year's holiday period which started in the week leading to 19/03/2020 and has the highest impact on reducing Rt.
†Older interventions end as soon as a new one takes into effect.
‡The relative reduction in Rt is based on estimates from European countries, USA, and Brazil.

**Fig. 5.**
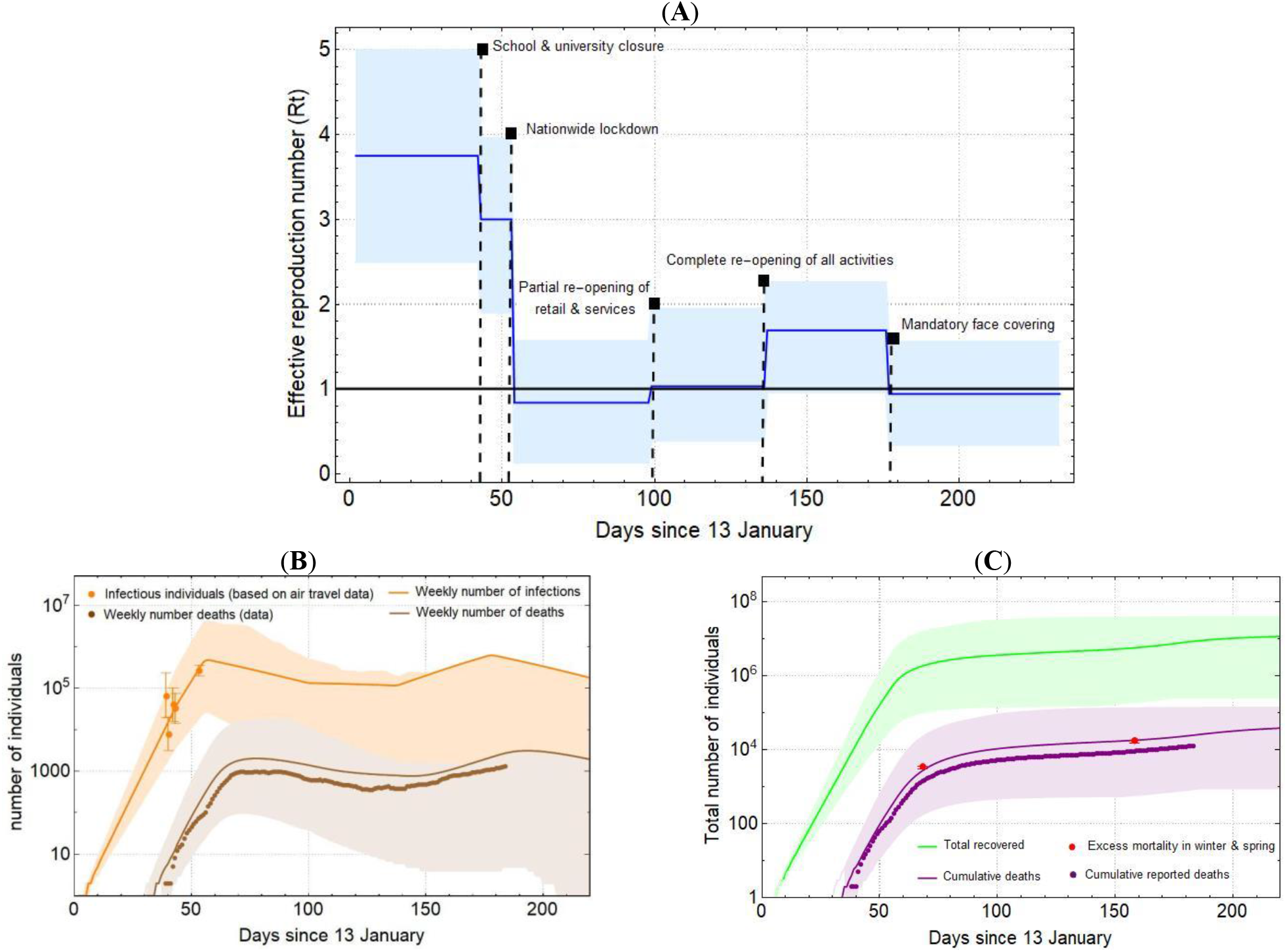
Reconstructing the epidemic in Iran using an SEIR model. Country-level estimation of infections, deaths, and proportion recovered based on the effect of NPIs on effective reproduction number. (**A**) Time-varying reproduction number, Rt. Vertical dashed lines correspond to the dates when major intervention measures were implemented (see Table 1). (**B**) Shows the weekly number of infections (orange line) and estimated number of infections (orange dots) based on confirmed exported cases (with error bars showing the 95% CI). The number of weekly infections estimated by the model drops immediately after an NPI takes into effect. The weekly number of deaths (brown line) follows the same trend as the reported number of deaths (brown dots). (**C**) Shows the cumulative deaths (purple line) and total number of individuals recovered (green line). The predicted cumulative deaths grow proportionally with respect to cumulative reported deaths (purple dots) and does not reach higher values with respect to the estimated excess mortality during winter and spring (red dots). Lines represent model predictions and shaded areas are 95% confidence intervals.

## Materials and Methods

### Data on aggregate number of cases and deaths

We use the time series data for the number of confirmed cases and deaths (see Data S2) from the John Hopkins University Centre for Systems Science and Engineering COVID-19 GitHub repository (accessed on 04/06/2020). We also obtain time series data on confirmed cases in all the 31 provinces of Iran from the ministry of health website (*behdasht.gov.ir*). We note that ministry stopped releasing province data from Mar 23 onward. They also did not release province data on Mar 2 and 3. We obtain mortality statistics from the National Organization for Civil Registration of Iran (*sabteahval.ir*).

### Benford’s law

Benford’s law (BL) defines a probability distribution for the leading digits in a determined set of numbers [27]. The probability that a digit, *d* = 1,2,…,9, is a leading number is given by *P*(*d*) = log_10_(1 + (1/ *d*)) where numbers with leading digit 1 have the highest probability of appearance and this probability steadily decreases as the starting digit becomes larger. BL is used in a variety of fields such as accounting, trade, and election results to study possible fraud and irregularities with data [28-30] and is also frequently used to assess the quality of epidemiological and clinical data [31-33].

In the early phase of an outbreak when the number of reported cases and deaths grow exponentially, a function of the form 2*^t^*^/^*^T^* exactly obeys the BL distribution, where *t* is the unit of time (measured in days) and *T* is the doubling time in new cases/deaths. If we combine two data sets with different growth rates into one larger data set, the numbers in the larger data set still obey the BL distribution. In the context of the COVID-19 outbreak, we can examine possible manipulation of data by comparing the probability distribution of the leading digits in the reported number of cases and deaths from different countries to BL distribution. We note that while this method can be used to test if data manipulation has occurred, it does not provide any information about deliberate absence of data by, for instance, not reporting deaths from certain hospitals.

### Phylogenetic analysis

We obtain the first whole-genome sequence of SARS-CoV-2 from inside Iran and collect an additional 20 sequences from cases that flew out of Iran and had their viral genomes determined in other countries. Two of these sequences are epidemiologically linked and cannot be used for the purpose of join-inference of TMRCA and doubling times. Thus, we use the remaining 19 whole-genome sequences to infer parameters of the epidemic using the phylogenetic software, BEAST [34]. We use the known sampling times with a Continuous-Time Markov Chain reference prior on the substitution rate, an exponential population coalescent model with a lognormal prior on size (mean=1, SD=2), Laplace prior on growth rate (scale=100), and an HKY+G substitution mode [23,24] and allow the Markov chain Monte Carlo to run for a hundred million steps and discard the first 10% steps as burn-in. Effective sample sizes of all parameters are >1,000 ensuring that they are well-sampled. We also construct the maximum likelihood tree of all sequences using PhyML [35].

### Estimating the number of active cases using air travel data

We estimate the number of passengers from four of the busiest airports in Iran (Tehran, Mashhad, Isfahan, and Shiraz) who flew to Oman, Lebanon, UAE, Kuwait, and China (see Data S3). We allow for 50% (20% - 100%) of the 55 million residents in provinces near the airports to represent their catchment population size (see Data S4). There is typically 5 (4-6) days of incubation period [7-9,36,37] and an additional 5 (3-7) days of delay from symptom onset to detection [38,39]. We further assume that an additional 45% (30%-55%) of asymptomatic and 15% (10%-25%) mildly-symptomatic cases were among the confirmed exported carriers of COVID-19 [40,41]. Thus, the probability of having an infected individual on board the aircraft is given by *p = Dt*/*M*, where *D* is the daily passenger flux, *M* is the catchment population size, and *t* is the exposure time which is the sum of incubation period and delay to symptom onset to detection. Finally, to estimate the number of infectious individuals,*λ*_1_, for country, *i*, with *n_i_* detected cases and probability of successful detection, *pi*, we can maximise the likelihood function given by

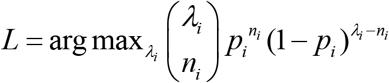

### Estimating seasonal excess mortality

We use the publicly available data from the NOCR which records the all-cause registered deaths in Iran per province per season in Solar Hijri (SH) calendar [42]. We assembled this data for the last 5 years to compare the excess mortality during winter and spring 2020 to previous years. Excess deaths refer to the number of deaths above expected seasonal baseline levels, regardless of the reported cause of death, and can be used as a nonspecific measure of the severity of the epidemic and provide a more accurate measure of its burden on the healthcare system [43]. We analyse the NOCR data from 1394 SH to 1399 SH (from 22 December 2015 to 20 June 2020 in the Gregorian calendar). For each province, we apply a least-square regression model on the seasonal deaths from previous years to find the ‘expected’ seasonal death in 2020 (see Fig. S1 and S2) and then calculate the difference with respect to the observed death toll for each province to find excess mortality (see Table S2 and S3). We attribute excess deaths in those provinces with significant deviations (i.e. three standard deviation units) with respect to their expected seasonal value to COVID-19-related deaths.

### Evaluating the importation risk of new cases from China in January

We define the importation risk as the mean number of infectious individuals travelling from China to Iran over the span of approximately three months, from the start of the outbreak in China until mid-March. We assume importations to Iran only occur via air travel from infected areas in China. The main routes of travel to Iran are from airports in Beijing, Shanghai, and Guangdong. We allow a fraction, *f*, of passengers to come from the Hubei province. To calculate *f* we take the ratio of total passenger traffic from airports in Hubei to 100 busiest airports across the country according to the 2018 report from the Civil Aviation Administration of China [44]. We further assume that only pre-symptomatic or asymptomatic cases travelled to Iran. The probability that an individual on board the plane is infected can be given by

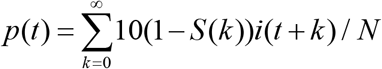

where *i*(*t*) is the incidence of cases in a given province at time *t*, *N* is the population size of the province, and *S*(*k*) is the CDF of the incubation period and is assumed to be log-normally distributed (mean=4.8 days, SD=1.9 days) [37]. The factor 10 is to correct for the true incidence by accounting for an additional 90% unascertained infections before mid-March in China [45]. Finally, to calculate the expected number of imported cases from 22 January to 15 March, we have

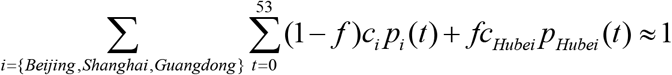

where *t*=0 corresponds to 22 January and *c_i_* is the average number of passengers flying from province *i* per day.

### SEIR model

We use a generalised SEIR model with age-stratified compartments to approximate the dynamics of the epidemic [46]. In this model, susceptible individuals can be exposed to the virus through contact with an infected individual. They then progress towards the infectious stage where they can either recover without hospitalisation or progress towards sever illness that requires hospitalisation. For the latter group, individuals either recover or transition to the ICU stage at which point they either die or return back to the hospitalisation state. The effective reproduction number, Rt, is modelled as a piecewise constant function that changes immediately upon a new NPI. In the absence of mobility data from Iran, we make the assumption that the efficacy of individual interventions from other countries are the same as those implemented in Iran and that they remain constant over time (see Table 2). Other studies have shown broad equivalency in the effect of reductions in different types of mobility and their corresponding impact on Rt [47-49]. We note that the dynamics of the epidemic, like any other exponentially growing process, is most sensitive to the growth rate. Thus, we allow for a relatively broad range in basic reproduction number, R_0_, and the efficacy of NPIs to appropriately capture the uncertainties in the dynamics. Furthermore, the model relies on fixed estimates of some epidemiological parameters such as age-stratified infection fatality ratio, latency period, and infectious period based on other studies (see Table 3). The duration of hospital/ICU stay is taken from the 14 March report by MoHME (see Fig. S3) [50], and the simulation start date is based on our analysis of exported cases (see Fig. 3b). We assume all infected individuals are immune to reinfection over the course of the simulation.

**Table 3.** SEIR model parameters. We use this information, along with the effectiveness of intervention measures in reducing Rt (Table 1), to run the simulations on *covid19-scenarios.org* dashboard [46].

| Age group | Age distribution* | % of all infections that are fatal† | Model parameters | Expected value | Reference |
| --- | --- | --- | --- | --- | --- |
| 0-9 | 14,434,000 | 0.00075 | Basic reproduction number | 3.75 (2.5-5) | [47-49,52] |
| 10-19 | 11,812,000 | 0.0045 | Latency period‡ | 2.5 days | [36,40,41] |
| 20-29 | 12,254,000 | 0.009 | Infectious period‡ | 4 days | [36,40,41] |
| 30-39 | 16,786,000 | 0.02 | Hospital stay | 4.5 days | Fig. S3 |
| 40-49 | 11,926,000 | 0.072 | ICU stay | 6 days | Fig. S3 |
| 50-59 | 8,281,000 | 0.25 | Imports per day | 0 |  |
| 60-69 | 5,264,000 | 1.1 | Initial number of cases | 1 |  |
| 70-79 | 2,264,000 | 3.1 | Simulation start date | 13/01/2020 |  |
| +80 | 1,015,000 | 6.9 | Simulation end date | 31/08/2020 |  |

\*based on annual reports on vital statistics from the Statistical Center of Iran (*amar.org.ir*).
†We use the statistics from the Chinese CDC [54] for age-stratified IFR which are also broadly compatible with estimates from other studies [55].
‡Latency period is the delay from infection to onset of infectiousness. Infectiousness typically starts from 2.5 days and peaks at 1 day before the onset of symptoms. Together with infectious period, latency period sets the serial interval [46].

## Discussion

Using various clinical, epidemiological, and genetic information, we reconstructed the outbreak of SARS-CoV-2 in Iran in the presence of significant levels of under-reporting of prevalence and deaths. We showed that Benford’s law can be used as a statistical test to investigate possible manipulation of reported data during the exponential phase of the epidemic. We then examined the burden of the outbreak on 31 provinces of Iran during winter and spring 2020 using the seasonal excess mortality data. We find that a large number of provinces did not show signals of significant excess mortality in winter further supporting the observation that the outbreak was still at its early stages during this period with only a few hot-spot provinces. We further showed that air travel data from confirmed exported cases acts as a proxy for measuring the number of active cases and that genomic data have the power to provide sensible estimates for the start date of the epidemic and its early doubling times. Phylogenetic analysis, coupled with epidemiological (recent travel to Iran in late February and early March) and clinical data (date of symptom onset), further helped with identification an emergent clade of SARS-CoV-2 linked to cases with a travel history to Iran. The lack of genetic diversity in these samples reflects the early stages of SARS-CoV-2 transmissions within Iran and newly identified sequences from Iran are likely to fall in this clade. Other phylogenetic analyses have shown that these sequences are also closely related to three Chinese strains sampled in mid-January from Hubei, Shandong, and Guangdong (Wuhan/HBCDC-HB-05/2020, Shandong/IVDC-SD-001/2020, and Guangdong/DG-S41-P0056/2020) indicating that the first imported case to Iran likely came from mainland China [21,22]. The overall low importation rate of new cases from China based on incidence data suggests that importation to Iran likely occurred via a high-risk individual with frequent travels to the country.

In this study, we found no significant evidence of data manipulations based on Benford’s law. However, this test alone cannot be used to rule out some systematic or random patterns of absence of data on case or death counts, e.g. from certain hospitals. The number of confirmed COVID-19 cases in each country may vary depending on the transparency to report correct statistics and the capacity of the healthcare system to detect new cases. The latter also depends on the accuracy of laboratory test kits and accessibility of diagnostic and screening tests. Indeed, many countries with limited testing capacities may have to prioritise testing that informs policy decisions, e.g. they may not test suspected cases with mild symptoms or those who are asymptomatic and are a close contact of a confirmed case. As a result, the true number of cases is always many times higher than those reported by the health agencies. In addition, the burden of the epidemic can significantly impact the performance of the healthcare system to properly allocate cause of deaths in individuals with underlying health conditions such as diabetes or heart disease. Therefore, tracking all-cause registered deaths and estimating excess mortality during the outbreak provides a more sensitive measure of COVID-19-associated deaths than would be recorded by counting confirmed or suspect deaths. We note that all-cause deaths may also include factors that are not causally associated with SARS-CoV-2 that might affect death rates such as the circulation of the 2019–20 seasonal influenza. According to the United Nations Statistical Division the estimated coverage of registered deaths in Iran is 92% [51] which could potentially be suffering even more from under-counting during the peak of the outbreak when there are likely going to be further delays in death registrations. Also, excess seasonal mortality does not take into account the catalysing role of COVID-19 in deaths among individuals with underlying comorbidities who would have likely died during a particular season even without contracting COVID-19 (as part of the ‘background’ deaths).

Despite integrating multiple sources, our data have many limitations. We investigated air travel data only to countries with direct flights to Iran and discarded information from detected cases in countries such as Qatar and Canada since we were not able to independently verify the fraction of passengers on board the planes who travel from Iran to those countries. Also, given the lack of mobility data from Iran, we were unable to investigate possible international exportation of cases to Afghanistan, Iraq, Syria, Azerbaijan, Turkey, and other countries with a significant flow of ground transportation (i.e. trains, buses, and cars) from Iran. We did not have access to the province-level number of confirmed COVID-19 deaths which is a significant source of information to assess excess deaths in the winter and spring 2020. Also, there is likely extreme heterogeneity in the geographical spread of COVID-19 across the country due to various factors such as demographic structure of the provinces’ populations, the pattern of social contacts between age groups, and the quality of healthcare and effectiveness of NPIs in different districts.

Our SEIR modelling analysis shows that in the most likely scenario by mid-July only 11% of the population have recovered from the disease which implies that a large fraction of the population is still vulnerable to contracting COVID-19. We also find strong indications that the outbreak was never brought under control since it emerged back in mid-January. This has significant implications both for the likely chance of the virus becoming endemic to the country and its likely return during winter this year which, if coupled with seasonal flu, can significantly overwhelm the hospitals. Furthermore, the continuation of under-reporting in prevalence due to limited testing of suspect cases and tracing their contacts will likely lead to several undetected superspreading events that can spark new outbreaks in different parts of the country making nowcasting and forecasting of the COVID-19 epidemic in Iran extremely challenging.

## Data Availability

We enclosed a supplementary file containing all data referred to in this manuscript.

## Funding

MG and SD are funded by the Biotechnology and Biological Science Research Council (BBSRC), grant number BB/M011224/1.

### Author contributions

MG, AL, and AK conceptualised and developed the work. MG wrote the manuscript and all other authors reviewed and edited the manuscript. BH and MG investigates data manipulation and carried out the Benford analysis. AKar collected the air travel data for confirmed exported cases. AKar and MG analysed the air travel data to measure incidence across the country. AKad and MG collected and analysed the excess mortality data. MAK, MA, and SZ collected and validated the genomic sample from Iran. AK and MG collected the remaining 20 genomic sequences from passengers with a travel history to Iran and carried out the phylogenetics analysis. MH and MG collected the data regarding daily reports on testing and set dates for non-pharmaceutical intervention measures.

### Data and materials availability

All data and codes used for the analysis is available online on our GitHub repository (github.com/mg878/Iran_study).

**Movies S1. The spread of the COVID-19 epidemic in Iran**.

This includes the incidence, total reported cases and deaths in 31 provinces from 19 February to 22 March 2020 (we note that MoHME has not released the province-level data on cases and deaths since 22 March).

**Data S1. Genome metadata**.

We gratefully acknowledge the authors, originating and submitting labs of the sequences from GISAID’s Database *(gisaid.org)* on which part of our genetic analysis is based.

**Data S2. Number of confirmed cases and deaths**.

This includes a time series data for the number of confirmed cases and deaths in Iran, UK, US, France, Italy, and Spain as well as daily confirmed cases in Guangdong, Shanghai, Beijing, and Hubei.

**Data S3. Estimated number of passengers per country per airport**.

This includes a detailed list of all flights from airports in Tehran, Isfahan, Shiraz, and Mashhad to Oman, UAE, Kuwait, Lebanon, and China. The list also includes flight numbers which enables us to find the approximate seating capacity based on aircraft model.

**Data S4. Estimated catchment population size**.

This includes the approximate population of 17 provinces that use the 4 airports in Iran for international travels.

**Data S5. SEIR simulation results**.

This includes a detailed output of our SEIR modelling (exported from *covid19-scenarios.org* dashboard). The file includes dates, confirmed weekly cases, confirmed weekly deaths, effective reproduction number, severe cases, exposed individuals, infectious individuals, severely ill individuals (hospitalised), individuals in the ICU, weekly fatalities, cumulative recovered, and cumulative fatalities with their corresponding 95% CI (upper and lower bounds).

**Fig. S1.**
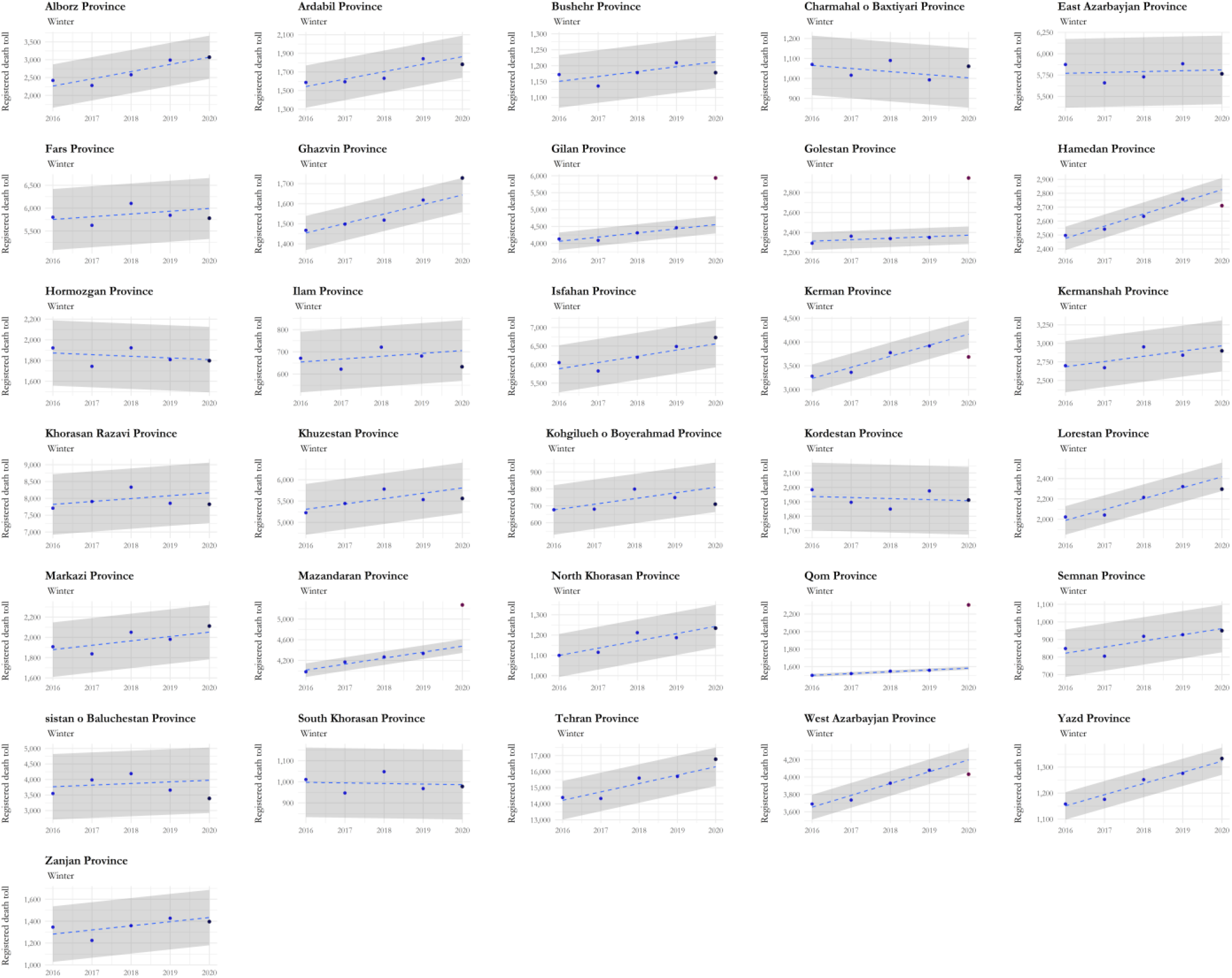
Province-level pattern of excess mortality during winter. This record covers every registered death over the last five years including last winter (from 22 December 2019 to 19 March 2020). Gray areas show the 95% confidence interval.

**Fig. S2.**
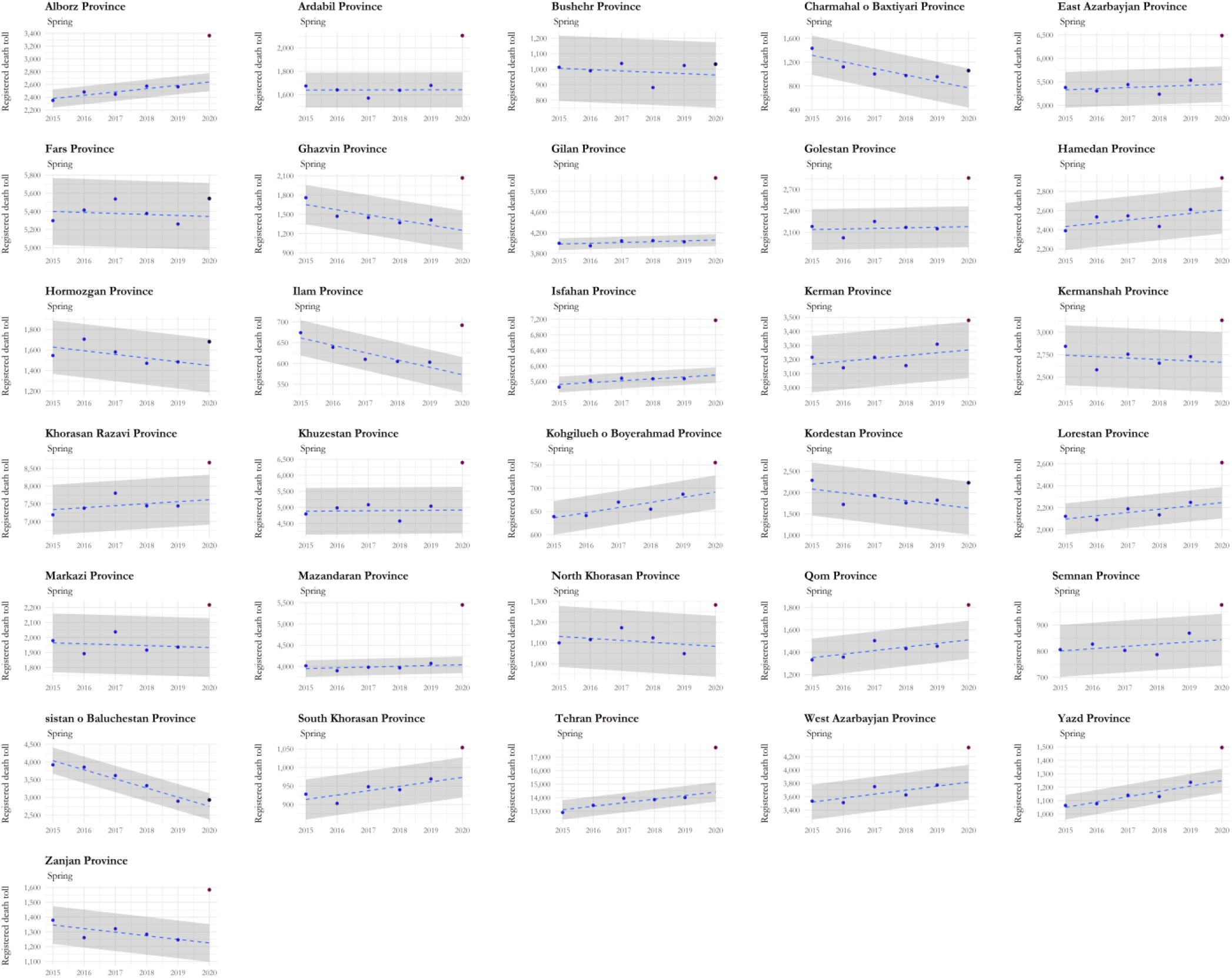
Province-level pattern of excess mortality during spring. This record covers every registered death over the last five years including last spring (from 20 March to 20 June 2020). Gray areas show the 95% confidence interval.

**Fig. S3.**
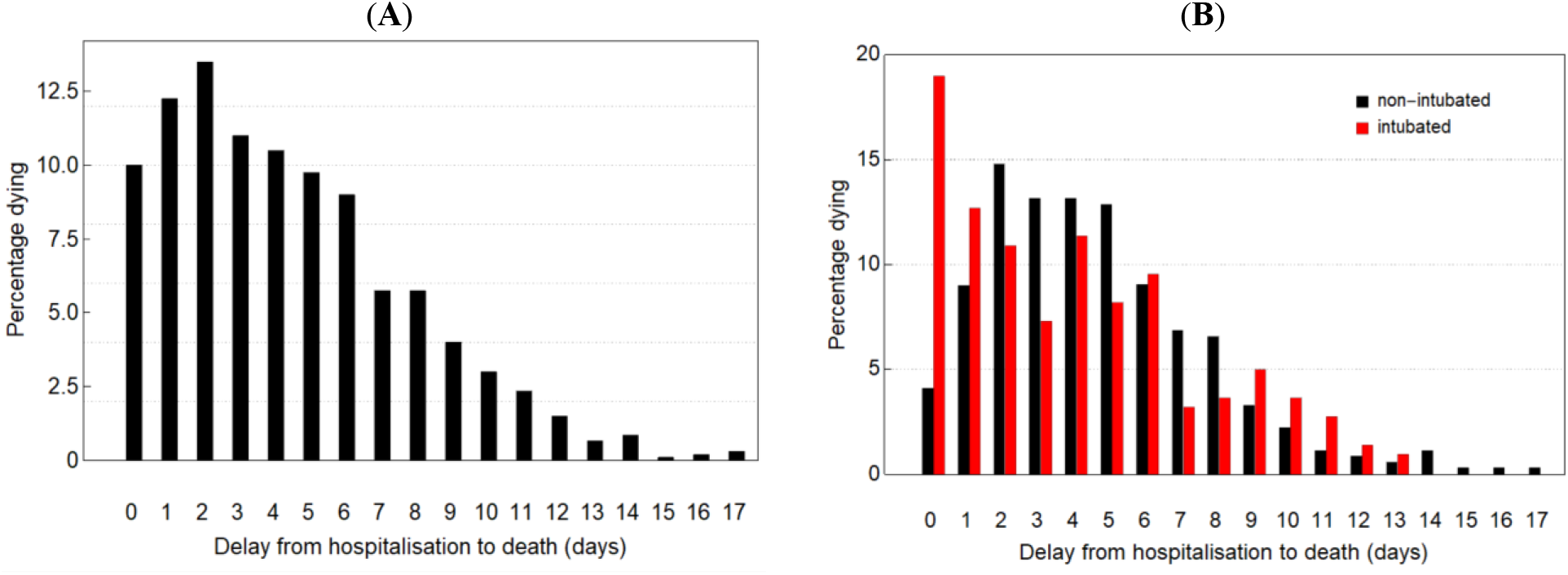
Delay from hospitalization to death in Iran. (**A**) Delay from hospital admission to hospital death [50]. (**B**) Comparison of delay from hospitalization to hospital death for patients with (red) and without tracheal intubation (black).

**Table S1.**
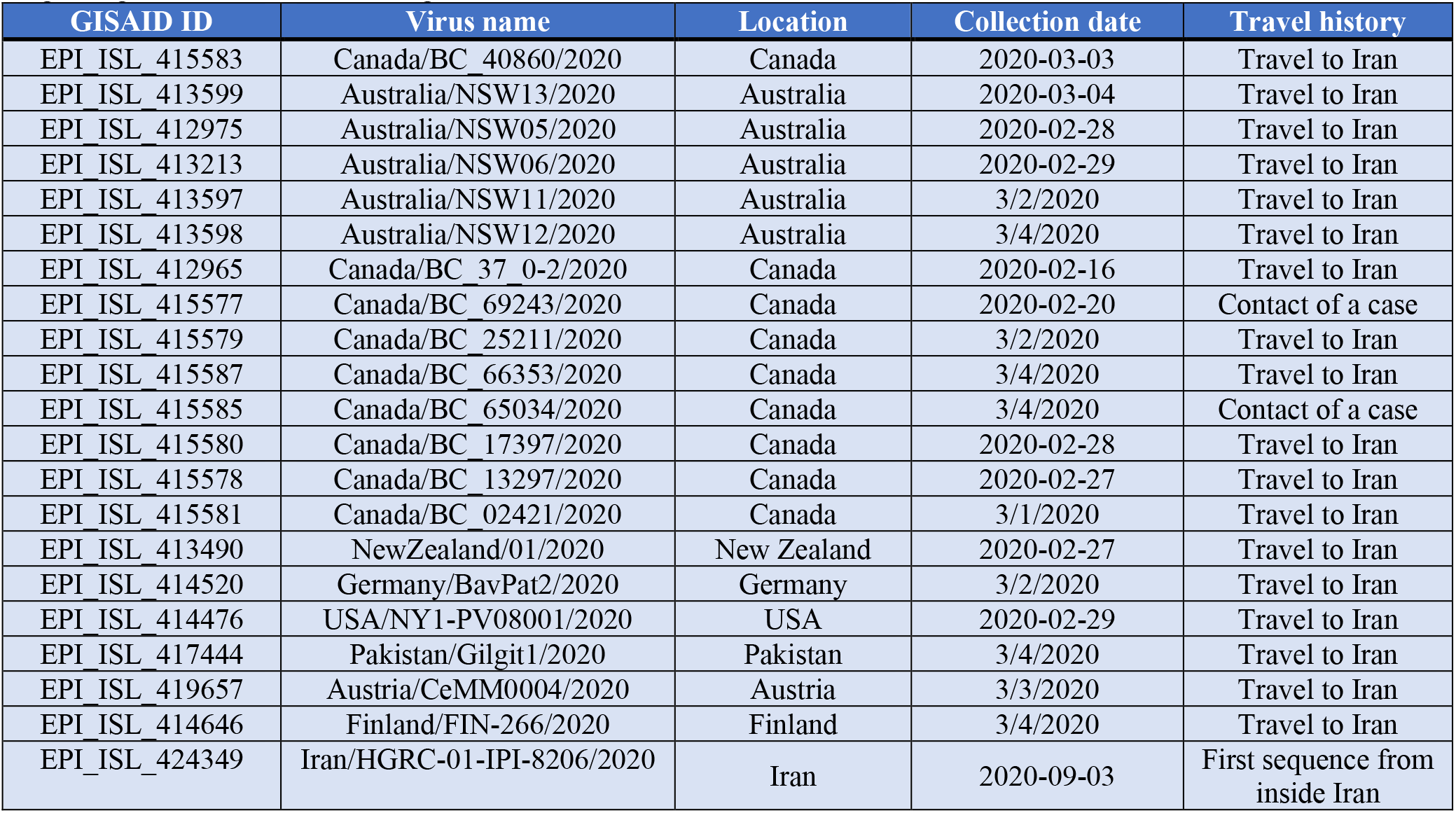
SARS-CoV-2 genomes analysed in this study. A complete metadata with additional information regarding author list and submitting labs can be found in Data S1.

| GISAID ID | Virus name | Location | Collection date | Travel history |
| --- | --- | --- | --- | --- |
| EPI_ISL_415583 | Canada/BC_40860/2020 | Canada | 2020-03-03 | Travel to Iran |
| EPI_ISL_413599 | Australia/NSW13/2020 | Australia | 2020-03-04 | Travel to Iran |
| EPI_ISL_412975 | Australia/NSW05/2020 | Australia | 2020-02-28 | Travel to Iran |
| EPI_ISL_413213 | Australia/NSW06/2020 | Australia | 2020-02-29 | Travel to Iran |
| EPI_ISL_413597 | Australia/NSW11/2020 | Australia | 3/2/2020 | Travel to Iran |
| EPI_ISL_413598 | Australia/NSW12/2020 | Australia | 3/4/2020 | Travel to Iran |
| EPI_ISL_412965 | Canada/BC_37_0-2/2020 | Canada | 2020-02-16 | Travel to Iran |
| EPI_ISL_415577 | Canada/BC_69243/2020 | Canada | 2020-02-20 | Contact of a case |
| EPI_ISL_415579 | Canada/BC_25211/2020 | Canada | 3/2/2020 | Travel to Iran |
| EPI_ISL_415587 | Canada/BC_66353/2020 | Canada | 3/4/2020 | Travel to Iran |
| EPI_ISL_415585 | Canada/BC_65034/2020 | Canada | 3/4/2020 | Contact of a case |
| EPI_ISL_415580 | Canada/BC_17397/2020 | Canada | 2020-02-28 | Travel to Iran |
| EPI_ISL_415578 | Canada/BC_13297/2020 | Canada | 2020-02-27 | Travel to Iran |
| EPI_ISL_415581 | Canada/BC_02421/2020 | Canada | 3/1/2020 | Travel to Iran |
| EPI_ISL_413490 | NewZealand/01/2020 | New Zealand | 2020-02-27 | Travel to Iran |
| EPI_ISL_414520 | Germany/BavPat2/2020 | Germany | 3/2/2020 | Travel to Iran |
| EPI_ISL_414476 | USA/NY1-PV08001/2020 | USA | 2020-02-29 | Travel to Iran |
| EPI_ISL_417444 | Pakistan/Gilgit1/2020 | Pakistan | 3/4/2020 | Travel to Iran |
| EPI_ISL_419657 | Austria/CeMM0004/2020 | Austria | 3/3/2020 | Travel to Iran |
| EPI_ISL_414646 | Finland/FIN-266/2020 | Finland | 3/4/2020 | Travel to Iran |
| EPI_ISL_424349 | Iran/HGRC-01-IPI-8206/2020 | Iran | 2020-09-03 | First sequence from inside Iran |

**Table S2.** Sensitivity analysis for the number of active cases based on confirmed exported cases.

| Lebanon (21 Feb) |  |  |  | Catchment population (M= million individuals) |  |  |  |
| --- | --- | --- | --- | --- | --- | --- | --- |
|  |  |  |  | min = 11.1M |  | max = 55.6M |  |
|  |  |  |  | Airplane capacity |  |  |  |
|  |  |  |  | min = 50% | max = 90% | min = 50% | max = 90% |
| Percentage of undetected pre- & asymptomatics | min =40% | Exposure time (days) | min = 7 | 54700 (16700 - 202900) | 30000 (8900 - 112300) | 277100 (87600 - 1018400) | 153500 (48200 - 565400) |
|  |  |  | max = 13 | 29000 (8600 - 108800) | 15700 (4300 - 60000) | 148800 (46700 - 547900) | 82300 (25500 - 304000) |
|  | max = 80% |  | min = 7 | 54700 (16700 - 202900) | 30000 (8900 - 112300) | 277100 (87600 - 1018400) | 153500 (48200 - 565400) |
|  |  |  | max = 13 | 29000 (8600 - 108800) | 15700 (4300 - 60000) | 148800 (46700 - 547900) | 82300 (25500 - 304000) |

| UAE (22 Feb) |  |  |  | Catchment population (M= million individuals) |  |  |  |
| --- | --- | --- | --- | --- | --- | --- | --- |
|  |  |  |  | min = 11.1M |  | max = 55.6M |  |
|  |  |  |  | Airplane capacity |  |  |  |
|  |  |  |  | min = 50% | max = 90% | min = 50% | max = 90% |
| Percentage of undetected pre- & asymptomatics | min =40% | Exposure time (days) | min = 7 | 5000 (1800 - 14600) | 2800 (1000 - 8000) | 24800 (9500 - 73200) | 13800 (5200 - 40600) |
|  |  |  | max = 13 | 2700 (900 - 7800) | 1500 (500 - 4300) | 13400 (5100 - 39400) | 7400 (2800 - 21800) |
|  | max = 80% |  | min = 7 | 6600 (2700 - 17000) | 3700 (1500 - 9400) | 33100 (14000 - 85500) | 18400 (7700 - 47400) |
|  |  |  | max = 13 | 3600 (1400 - 9100) | 2000 (1500 - 5000) | 17800 (7500 - 46000) | 9900 (4100 - 25500) |

| Oman (24 Feb) |  |  |  | Catchment population (M= million individuals) |  |  |  |
| --- | --- | --- | --- | --- | --- | --- | --- |
|  |  |  |  | min = 11.1M |  | max = 55.6M |  |
|  |  |  |  | Airplane capacity |  |  |  |
|  |  |  |  | min = 50% | max = 90% | min = 50% | max = 90% |
| Percentage of undetected pre- & asymptomatics | min =40% | Exposure time (days) | min = 7 | 25,100 (9,200 – 74,200) | 13,900 (5,100 – 41,200) | 125,400 (46,600 – 371,400) | 69,700 (25,800 – 206,300) |
|  |  |  | max = 13 | 13,500 (4,900 – 39,900) | 7,500 (2,700 – 22,100) | 67500 (25000 - 199900) | 37,500 (13,900 – 111,000) |
|  | max = 80% |  | min = 7 | 33,400 (13,800 – 86,600) | 18,600 (7,600 – 48,100) | 167200 (69200 - 433400) | 92,900 (38,400 – 240,700) |
|  |  |  | max = 13 | 18,000 (7,400 – 46,600) | 10,000 (4,000 – 25,800) | 90,000 (37,200 – 233,300) | 50,000 (20,600 – 129,600) |

| Kuwait (25 Feb) |  |  |  | Catchment population (M= million individuals) |  |  |  |
| --- | --- | --- | --- | --- | --- | --- | --- |
|  |  |  |  | min = 11.1M |  | max = 55.6M |  |
|  |  |  |  | Airplane capacity |  |  |  |
|  |  |  |  | min = 50% | max = 90% | min = 50% | max = 90% |
| Percentage of undetected pre- & asymptomatics | min =40% | Exposure time (days) | min = 7 | 5,000 (1,800 – 14,600) | 2,800 (1,000 – 8,000) | 24,800 (9,500 – 73,200) | 13,800 (5,200 – 40,600) |
|  |  |  | max = 13 | 2,700 (900 – 7,800) | 1,500 (500 – 4,300) | 13,400 (5,100 – 39,400) | 7,400 (2,800 – 21,800) |
|  | max = 80% |  | min = 7 | 6,600 (2,700 – 17,000) | 3,700 (1,500 – 9,400) | 33100 (14,000 – 85,500) | 18400 (7,700 – 47,400) |
|  |  |  | max = 13 | 3,600 (1,400 – 9,100) | 2,000 (1,500 – 5,000) | 17800 (7,500 – 46,000) | 9900 (4,100 – 25,500) |

| China (6 Mar) |  |  |  | Catchment population (M= million individuals) |  |  |  |
| --- | --- | --- | --- | --- | --- | --- | --- |
|  |  |  |  | min = 11.1M |  | max = 55.6M |  |
|  |  |  |  | Airplane capacity |  |  |  |
|  |  |  |  | min = 50% | max = 90% | min = 50% | max = 90% |
| Percentage of undetected pre- & asymptomatics | min =40% | Exposure time (days) | min = 7 | 197,700 (149,600 – 277,000) | 109,800 (83,100 – 153,800) | 988,400 (748,400 – 1,385,300) | 549,100 (415,700 – 769,600) |
|  |  |  | max = 13 | 106,400 (80,500 – 149,100) | 59,100 (44,700 – 82,800) | 532200 (402900 - 745900) | 295,700 (223,800 – 414,300) |
|  | max = 80% |  | min = 7 | 252,100 (196,400 – 339,100) | 140,000 (109,100 – 188,400) | 1260300 (982300 - 1696100) | 700,100 (545,700 – 942,200) |
|  |  |  | max = 13 | 135,700 (105,700 – 182,600) | 75,400 (58,700 – 101,400) | 678,600 (528,900 – 91,3200) | 377,000 (293,800 – 507,300) |

**Table S2.**
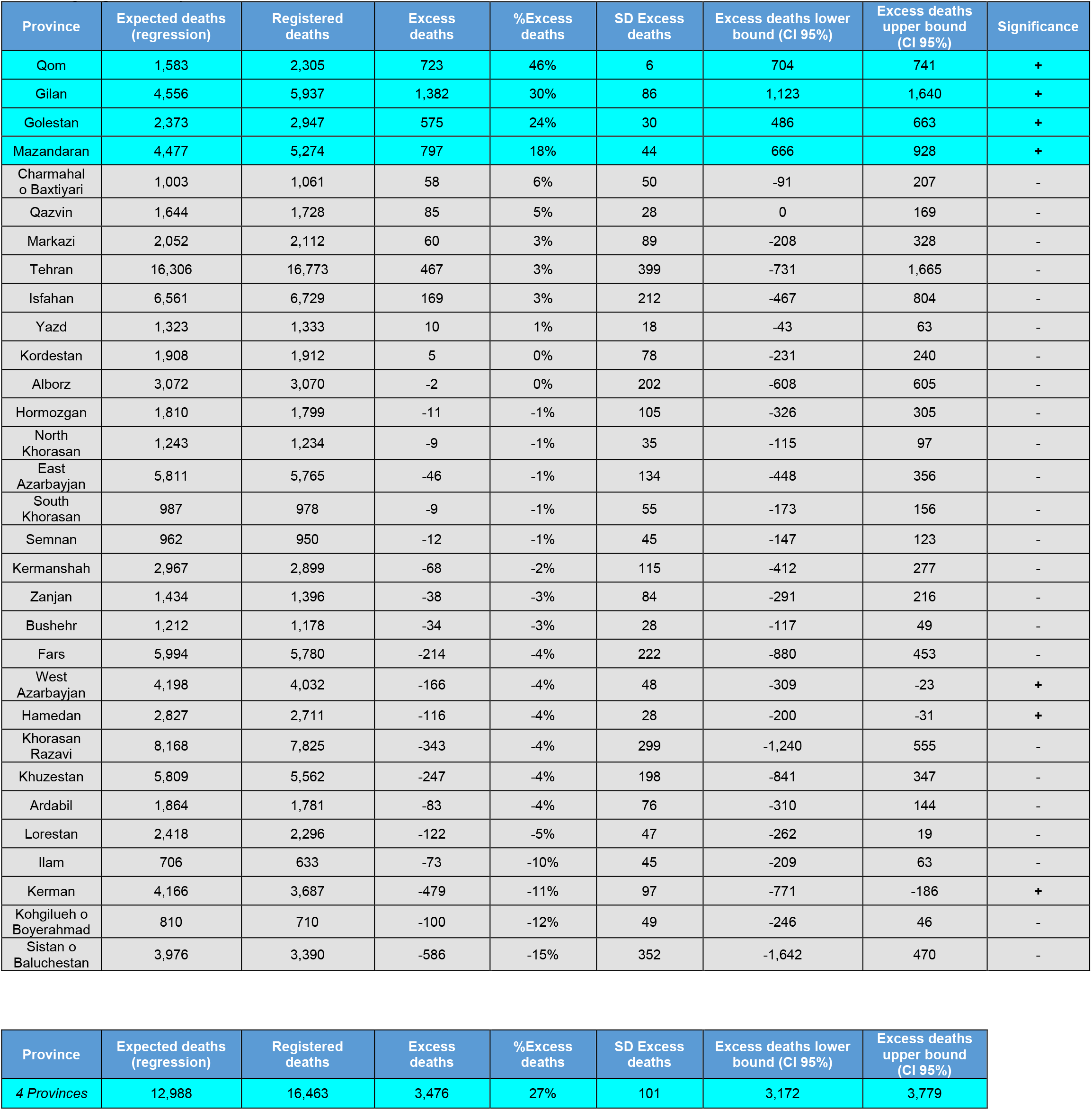
Excess mortality in 31 provinces during Winter 2020. This record covers every registered death from 22 December 2019 to 19 March 2020. Provinces with significantly higher excess deaths compared to previous years are highlighted in cyan.

| Province | Expected deaths (regression) | Registered deaths | Excess deaths | %Excess deaths | SD Excess deaths | Excess deaths lower bound (CI 95%) | Excess deaths upper bound (CI 95%) | Significance |
| --- | --- | --- | --- | --- | --- | --- | --- | --- |
| Qom | 1,583 | 2,305 | 723 | 46% | 6 | 704 | 741 | + |
| Gilan | 4,556 | 5,937 | 1,382 | 30% | 86 | 1,123 | 1,640 | + |
| Golestan | 2,373 | 2,947 | 575 | 24% | 30 | 486 | 663 | + |
| Mazandaran | 4,477 | 5,274 | 797 | 18% | 44 | 666 | 928 | + |
| Charmahal o Baxtiyari | 1,003 | 1,061 | 58 | 6% | 50 | -91 | 207 | - |
| Qazvin | 1,644 | 1,728 | 85 | 5% | 28 | 0 | 169 | - |
| Markazi | 2,052 | 2,112 | 60 | 3% | 89 | -208 | 328 | - |
| Tehran | 16,306 | 16,773 | 467 | 3% | 399 | -731 | 1,665 | - |
| Isfahan | 6,561 | 6,729 | 169 | 3% | 212 | -467 | 804 | - |
| Yazd | 1,323 | 1,333 | 10 | 1% | 18 | -43 | 63 | - |
| Kordestan | 1,908 | 1,912 | 5 | 0% | 78 | -231 | 240 | - |
| Alborz | 3,072 | 3,070 | -2 | 0% | 202 | -608 | 605 | - |
| Hormozgan | 1,810 | 1,799 | -11 | -1% | 105 | -326 | 305 | - |
| North Khorasan | 1,243 | 1,234 | -9 | -1% | 35 | -115 | 97 | - |
| East Azarbaijan | 5,811 | 5,765 | -46 | -1% | 134 | -448 | 356 | - |
| South Khorasan | 987 | 978 | -9 | -1% | 55 | -173 | 156 | - |
| Semnan | 962 | 950 | -12 | -1% | 45 | -147 | 123 | - |
| Kermanshah | 2,967 | 2,899 | -68 | -2% | 115 | -412 | 277 | - |
| Zanjan | 1,434 | 1,396 | -38 | -3% | 84 | -291 | 216 | - |
| Bushehr | 1,212 | 1,178 | -34 | -3% | 28 | -117 | 49 | - |
| Fars | 5,994 | 5,780 | -214 | -4% | 222 | -880 | 453 | - |
| West Azarbaijan | 4,198 | 4,032 | -166 | -4% | 48 | -309 | -23 | + |
| Hamedan | 2,827 | 2,711 | -116 | -4% | 28 | -200 | -31 | + |
| Khorasan Razavi | 8,168 | 7,825 | -343 | -4% | 299 | -1,240 | 555 | - |
| Khuzestan | 5,809 | 5,562 | -247 | -4% | 198 | -841 | 347 | - |
| Ardabil | 1,864 | 1,781 | -83 | -4% | 76 | -310 | 144 | - |
| Lorestan | 2,418 | 2,296 | -122 | -5% | 47 | -262 | 19 | - |
| Ilam | 706 | 633 | -73 | -10% | 45 | -209 | 63 | - |
| Kerman | 4,166 | 3,687 | -479 | -11% | 97 | -771 | -186 | + |
| Kohgilueh o Boyerahmad | 810 | 710 | -100 | -12% | 49 | -246 | 46 | - |
| Sistan o Baluchestan | 3,976 | 3,390 | -586 | -15% | 352 | -1,642 | 470 | - |

| Province | Expected deaths (regression) | Registered deaths | Excess deaths | %Excess deaths | SD Excess deaths | Excess deaths lower bound (CI 95%) | Excess deaths upper bound (CI 95%) |
| --- | --- | --- | --- | --- | --- | --- | --- |
| 4 Provinces | 12,988 | 16,463 | 3,476 | 27% | 101 | 3,172 | 3,779 |

**Table S3.** Excess mortality in 31 provinces during 2020. This record covers every registered death from 20 March to 20 June 2020. Provinces with significantly higher excess deaths compared to previous years are highlighted in green.

| Province | Expected deaths (regression) | Registered deaths | Excess deaths | %Excess deaths | SD Excess deaths | Excess deaths lower bound (CI 95%) | Excess deaths upper bound (CI 95%) | Significance |
| --- | --- | --- | --- | --- | --- | --- | --- | --- |
| Qazvin | 1,251 | 2,067 | 816 | 65% | 103 | 508 | 1,123 | + |
| Charmahal o Baxtiyari | 766 | 1,057 | 291 | 38% | 110 | -40 | 622 | - |
| Kordestan | 1,635 | 2,229 | 594 | 36% | 207 | -27 | 1,215 | - |
| Mazandaran | 4,040 | 5,449 | 1,409 | 35% | 65 | 1,214 | 1,604 | + |
| Golestan | 2,180 | 2,858 | 678 | 31% | 95 | 394 | 963 | + |
| Khuzestan | 4,919 | 6,395 | 1,476 | 30% | 242 | 751 | 2,201 | + |
| Gilan | 4,059 | 5,258 | 1,199 | 30% | 38 | 1,085 | 1,313 | + |
| Zanjan | 1,225 | 1,585 | 360 | 29% | 43 | 232 | 488 | + |
| Ardabil | 1,641 | 2,105 | 464 | 28% | 50 | 314 | 614 | + |
| Alborz | 2,638 | 3,365 | 727 | 28% | 47 | 585 | 869 | + |
| Isfahan | 5,767 | 7,172 | 1,405 | 24% | 67 | 1,204 | 1,607 | + |
| Tehran | 14,421 | 17,713 | 3,292 | 23% | 239 | 2,575 | 4,008 | + |
| Ilam | 573 | 692 | 119 | 21% | 14 | 76 | 161 | + |
| Qom | 1,511 | 1,823 | 312 | 21% | 57 | 141 | 483 | + |
| Yazd | 1,249 | 1,496 | 247 | 20% | 30 | 157 | 338 | + |
| East Azarbayjan | 5,451 | 6,485 | 1,034 | 19% | 126 | 655 | 1,414 | + |
| North Khorasan | 1,083 | 1,283 | 200 | 18% | 49 | 53 | 347 | + |
| Kermanshah | 2,666 | 3,132 | 466 | 17% | 111 | 133 | 799 | + |
| Lorestan | 2,246 | 2,611 | 365 | 16% | 47 | 223 | 507 | + |
| Hormozgan | 1,449 | 1,681 | 232 | 16% | 87 | -29 | 493 | - |
| Semnan | 844 | 977 | 133 | 16% | 33 | 34 | 232 | + |
| Markazi | 1,933 | 2,217 | 284 | 15% | 65 | 88 | 479 | + |
| Khorasan Razavi | 7,618 | 8,665 | 1,047 | 14% | 235 | 342 | 1,752 | + |
| West Azarbayjan | 3,818 | 4,338 | 520 | 14% | 87 | 258 | 781 | + |
| Hamedan | 2,605 | 2,940 | 335 | 13% | 82 | 90 | 579 | + |
| Kohgilueh o Boyerahmad | 691 | 755 | 64 | 9% | 12 | 28 | 100 | + |
| South Khorasan | 973 | 1,054 | 81 | 8% | 18 | 27 | 135 | + |
| Bushehr | 963 | 1,033 | 70 | 7% | 70 | -141 | 280 | - |
| Sistan o Baluchestan | 2,750 | 2,929 | 179 | 7% | 124 | -193 | 551 | - |
| Kerman | 3,268 | 3,479 | 211 | 6% | 67 | 11 | 411 | + |
| Fars | 5,343 | 5,541 | 198 | 4% | 123 | -172 | 567 | - |

| Province | Expected deaths (regression) | Registered deaths | Excess deaths | %Excess deaths | SD Excess deaths | Excess deaths lower bound (CI 95%) | Excess deaths upper bound (CI 95%) |
| --- | --- | --- | --- | --- | --- | --- | --- |
| 26 Provinces | 78,672 | 95,914 | 17,242 | 22% | 518 | 15,687 | 18,797 |

